# Early detection of in-patient deterioration: one prediction model does not fit all

**DOI:** 10.1101/2020.04.24.20077453

**Authors:** Jacob N. Blackwell, Jessica Keim-Malpass, Matthew T. Clark, Rebecca L. Kowalski, Salim N. Najjar, Jamieson M. Bourque, Douglas E. Lake, J. Randall Moorman

## Abstract

**Objectives:** Early detection of subacute potentially catastrophic illnesses using available data is a clinical imperative, and scores that report risk of imminent events in real time abound. Patients deteriorate for a variety of reasons, and it is unlikely that a single predictor such as an abnormal National Early Warning Score (NEWS) will detect all of them equally well. The objective of this study was to test the idea that the diversity of reasons for clinical deterioration leading to ICU transfer mandates multiple targeted predictive models.

**Design:** Individual chart review to determine the clinical reason for ICU transfer; determination of relative risks of individual vital signs, lab tests and cardiorespiratory monitoring measures for prediction of each clinical reason for ICU transfer; logistic regression modeling for the outcome of ICU transfer for a specific clinical reason.

**Setting:** Cardiac medical-surgical ward; tertiary care academic hospital.

**Patients:** 8111 adult patients, 457 of whom were transferred to an ICU for clinical deterioration.

**Interventions:** None.

**Measurements and main results:** We calculated the contributing relative risks of individual vital signs, lab tests and cardiorespiratory monitoring measures for prediction of each clinical reason for ICU transfer, and used logistic regression modeling to calculate ROC areas and relative risks for the outcome of ICU transfer for a specific clinical reason. The reasons for clinical deterioration leading to ICU transfer were varied, as were their predictors. For example, the three most common reasons – respiratory instability, infection and suspected sepsis, and heart failure requiring escalated therapy – had distinct signatures of illness. Statistical models trained to target specific reasons for ICU transfer performed better than one model targeting combined events, and both performed better than the untrained NEWS score.

**Conclusions and relevance:** A single predictive model for clinical deterioration does not perform as well as having multiple models trained for the individual specific clinical events leading to ICU transfer.

## Introduction

Patients who deteriorate on the hospital ward and are emergently transferred to the ICU have poor outcomes.^1–6^ Early identification of subtly worsening patients might allow quicker treatment and improved outcome. To aid clinicians, early warning scores have been advanced that compare vital signs and lab tests to thresholds. The reception has been mixed. ^7,8^ Vital signs are often delayed, incorrect, or never measured,^9,10^ and lab tests require blood draws and time. Nonetheless, these are the major (or only) inputs into popular track-and-trigger systems such as the National Early Warning Score (NEWS) and others.^11–20^ Thresholds for awarding risk points are based on clinical experience, but suffer from lack of validation and the loss of information that forced dichotomization inevitably brings.^21–24^ Only some are trained on clinical events. For example, eCART is trained for early detection of hospital patients with cardiac arrest ^14^, and the Rothman Index is trained to detect patients who will die in the next 12 months ^15^.

NEWS has been used widely as an early warning score of clinical deterioration since it was adopted in 2012 by the Royal College of Physicians and the UK National Health Service. A primary strength of general track-and-trigger systems like NEWS is in promoting inter-provider communication.^25–27^ On the other hand, a meta-analysis ^28^ of 232 studies emphasized multiple shortcomings of this kind of early warning tool, including the intermittent sampling, vulnerability to inaccuracies and user error, limited sensitivity, and lack of specialty-specific predictors.^29–32^

Importantly, most systems ignore the wealth of information that is present in cardiorespiratory monitoring. While many patients are continuously monitored, these data are bulky and require mathematical analysis. Nonetheless, they contribute equally to vital signs and lab tests in detection of patients at risk of clinical deterioration and ICU transfer from a cardiac medical and surgical ward. ^39^ We note, moreover, that risk estimation for neonatal sepsis using advanced mathematical analysis that considers only continuous cardiorespiratory monitoring saves lives.^33^

Finally, all of these early detection systems present one score as an omniscient risk marker for all conditions and patients.^34^ Clinicians know, though, that there are many paths to clinical deterioration. For example, the most common forms of deterioration leading to ICU transfer are respiratory instability,^35–41^ hemodynamic instability^35,38,39^, sepsis^40,42^, bleeding^38,42^, neurological decompensation^42^, unplanned surgery^38^, and acute renal failure or electrolyte abnormalities.^42^ The multiplicity of candidate culprit organ systems suggests that a single model is not likely to catch all the patients who are worsening.^38,39^

More recent evidence of the heterogeneity of human illness and the elusive nature of early detection lies in the results of the 2019 Computing in Cardiology Challenge.^43^ Contestants trained models on data from 2 hospitals and tested them on a held-out data from a third hospital. We note two important results. First, the best scores on the training data were on the order of 40 to 50% of the highest possible score. Second, the scores fell a great deal – most to negative numbers, signifying worse performance than an algorithm that never predicted sepsis – when tested in the third hospital. We take this as more evidence of the difficulty of detection of even a single medical diagnosis, sepsis, let alone all possible ones at the same time.

Here, we tested three approaches to predictive analytics monitoring. The first is untrained models that apply thresholds and cutoffs to vital signs, and we represent the class with NEWS. The second was to train a universal predictive model on all patients who went to the ICU using measured values of vital signs, lab tests and continuous cardiorespiratory monitoring. This approach has advantages of learning from continuous cardiorespiratory monitoring and from the range of measured vital signs and lab test values, avoiding problems of dichotomization by thresholds. The third is a set of models trained on patients who had specific reasons for ICU transfer identified by clinician review, which has the additional advantage of learning signatures of specific target illnesses.

Approaches that might improve track-and-trigger systems include the use of libraries of predictive models that are tailored for specific reasons for clinical deterioration within the widely varied venues within the modern hospital, risk estimation along the continuum of measured values rather than threshold crossings, and analysis of continuous data in addition to vital signs and lab tests. Here we test these ideas.

## Methods

### Study design

In early detection of hospital patients at risk of clinical deterioration and the need of escalation to ICU care, we wished to test the idea that a single predictive analytics model using thresholds of vital signs and lab tests could be improved upon by a set of target-specific models that used continuous risk estimates and cardiorespiratory monitoring.

We examined the empiric risk profiles of individual vital signs, lab tests and cardiorespiratory monitoring parameters for 7 clinical deterioration phenotypes. We made predictive statistical models based on logistic regression adjusted for repeated measures for each phenotype as well as their composite, and examined their performance in detecting other deterioration phenotypes. The University of Virginia Institutional Review Board approved the study.

### Study population

We studied 8111 consecutive admissions from October 2013 to September 2015 on a 73-bed adult acute care cardiac and cardiovascular surgery ward at the University of Virginia Hospital.^39^ We used an institutional electronic data warehouse to access electronic medical record (EMR). Six patients were added to the original cohort (n= 8105) who did not have complete continuous cardiorespiratory monitoring data.

We reviewed the charts of the 457 patients who were transferred to the ICU because of clinical deterioration. Five clinical reviewers developed and implemented clinical definitions indicating reasons for deterioration and reviewed records for the 48 hours prior to and following ICU transfer. To evaluate inter-rater reliability, we calculated a weighted kappa coefficient on a nested 50 events that were evaluated by all reviewers. The new clinical review was independent of our earlier work, ^39^ which used a combined outcome of ICU transfer, urgent surgery, or unexpected death.

### Clinical and continuous cardiorespiratory monitoring data

We analyzed vital signs and laboratory tests recorded in the EMR, and the continuous 7-lead EKG signal that is standard of practice. Vital signs (Figure 3) and laboratory tests (Figure 4) were sampled and held. We calculated cardiorespiratory dynamics at 15-minute intervals over 30-minute windows.

### Statistical analyses

To estimate the relative risk of an event as a function of measured variables, we constructed predictiveness curves ^44^. In order to reduce bias due to repeated measures and missing data, we used a bootstrapping technique. We sampled one measurement within 12 hours before each event and one measurement from all non-event patients at a random time during their stay. The distributions of time since admission were not significantly different for these two groups (data not shown). We calculated the relative risk of an event at each decile of the sampled variable, then interpolated the risk to 20 points evenly spaced in the range of the variable. We repeated this process of sampling, calculating relative risk, and interpolating 30 times. Finally, we averaged the 30 risk estimate curves to obtain a bootstrapped predictiveness curve at the 20 evenly spaced points, and we display the result as a heat map.

To relate the predictors to the outcomes, we used multivariable logistic regression analysis adjusted for repeated measures. We labelled data within 12 hours of ICU transfer as outcome=1 and the rest as outcome=0. The output of the resulting regression expressions is the probability of ICU transfer in the next 12 hours.

To test for the significance of differences between the outputs of predictive models, we calculated the area under the receiver operating characteristic (ROC) curves for NEWS, a model trained to target all the ICU transfers, and 7 additional models, one for each reason for ICU transfer. The ROC curves report on discrimination between data belonging to patients with and without ICU transfer in the next 12 hours. For each reason for transfer, therefore, we calculated 3 ROC areas. For the group of 7 specific reasons for ICU transfer, we tested the difference between NEWS and the outputs of the general model, and between the outputs of the general model and the outputs of the 7 specifically targeted models using the paired t-test.

In model development, we adhered to the Transparent Reporting of multivariable prediction model for Individual Prognosis Or Diagnosis (TRIPOD) statement recommendations.^45^

## Results

### Study population

The main admitting services were cardiology (49%) and cardiac surgery (19%); the remainder were equally from other medical or surgical services. The median age was 59 (IQR 55 to 75). The mortality rate was 0.4% and 17% for patients who did not or did require ICU transfer.

Figure 1 is an UpSet plot^46^ of the numbers of patients and their reasons for ICU transfer. The most common reason for transfer was respiratory instability alone, followed by respiratory instability and suspected sepsis. The kappa statistic ranged from 0.616 to 1.0 indicating moderate to excellent agreement among the reviewers.

**Figure 1.**
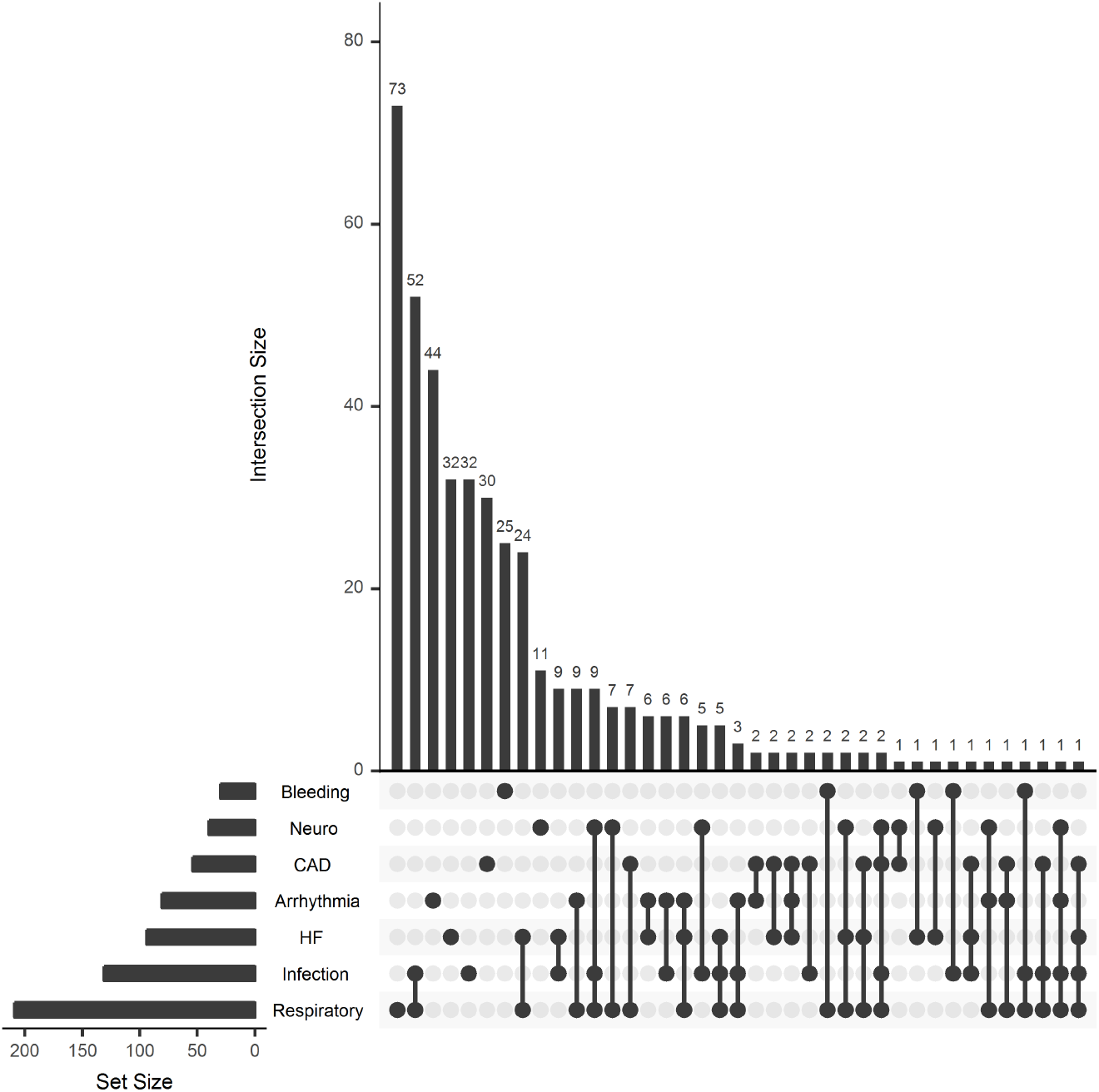
Reasons for ICU transfer. The horizontal bars show the number of patients in each category of the reason for ICU transfer; the vertical bars show the numbers of patients arranged by the presence of one or more reasons. The classifications were respiratory instability (increase in oxygen requirement, oxygen desaturation, respiratory distress or increased work of breathing); infection workup (culture taken and antibiotics started or broadened, proven infection that could have reasonably contributed to the deterioration); heart failure exacerbation (pulmonary edema, persistent hypotension, poor diuresis, severe pulmonary hypertension); arrhythmia (ventricular or supraventricular tachyarrhythmia, bradyarrhythmia); unstable coronary artery disease (unstable angina, acute myocardial infarction with or without ST segment elevation); neurological event (stroke, seizure, or altered mental status); and bleeding (transfusion requirement, hematoma, post-surgical bleed, GI bleed). Event classifications were not mutually exclusive, and many patients met more than one classification.

### Relative risks for ICU admission

Figure 2 shows the relationship of risk (color) to reason for admission to ICU (row) and measured variable (column). Each of the three panels shows a set of predictiveness curves as heat maps for kinds of measurements – a lab test (Na) in panel A, a vital sign (mean arterial pressure, MAP) in panel B, and a measure from continuous cardiorespiratory monitoring (breathing rate derived from the EKG^47^ in panel C. The gray-scale bars at the top show the densities of the measurements.

**Figure 2.**
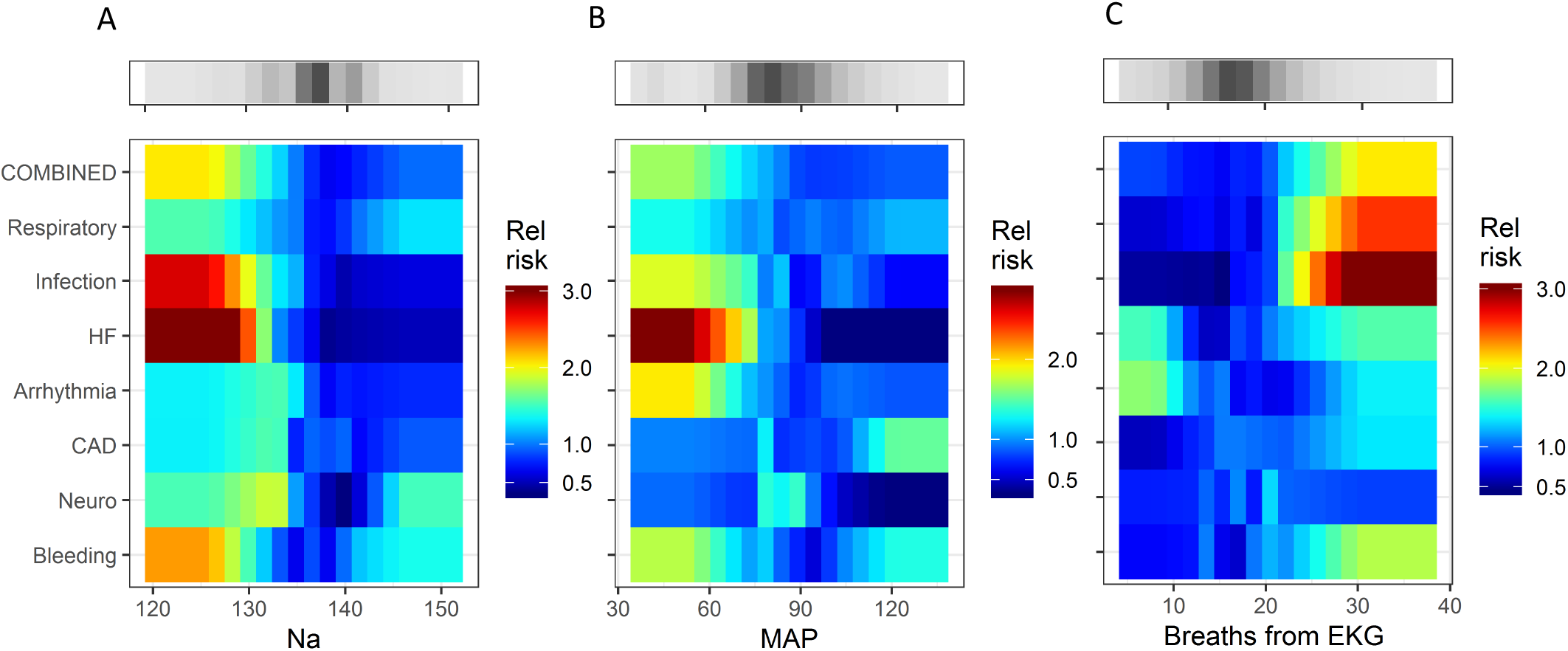
Heat maps that relate the relative risk (color bar; 1 means the risk is average for the ward) to the reason for ICU transfer (rows) and measured value of a predictor (abscissa) for a representative laboratory value (A, serum Na in meq/dl), vital sign (B, mean arterial pressure, MAP in mm Hg), and cardiorespiratory measure from time series analysis of continuous cardiorespiratory monitoring (C, breaths per minute derived from the EKG). The gray-scale bars show the density of the measurements. The Figure may be interpreted as follows. The change in color from deep red to blue from left to right in the fourth row of panel A signifies that low serum Na concentration was associated with a high risk for ICU transfer for escalation of heart failure (HF) therapy.

**Figure 3.**
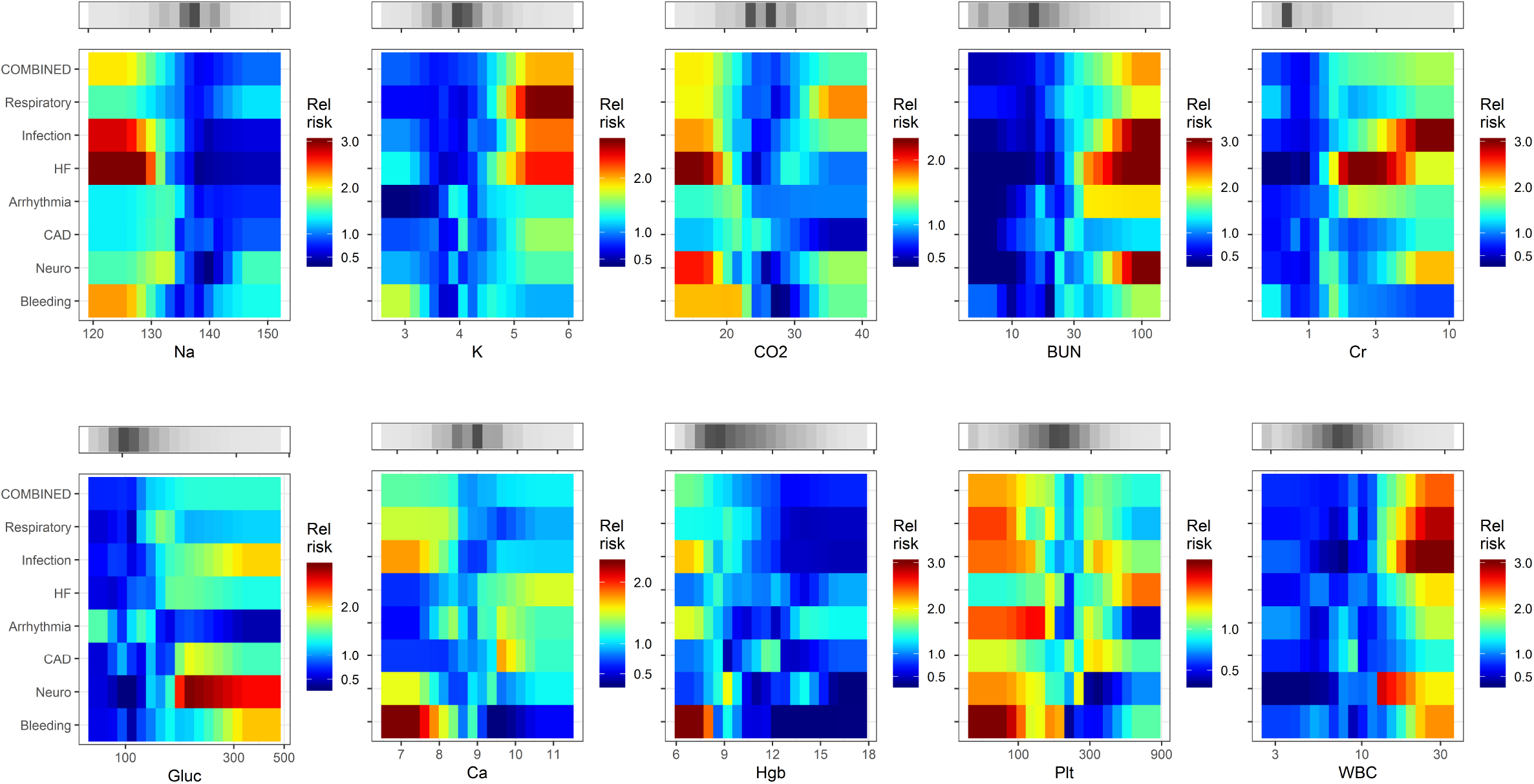
Heat maps that relate the relative risk of the reason for ICU transfer to the measured values of predictor laboratory values.

**Figure 4.**
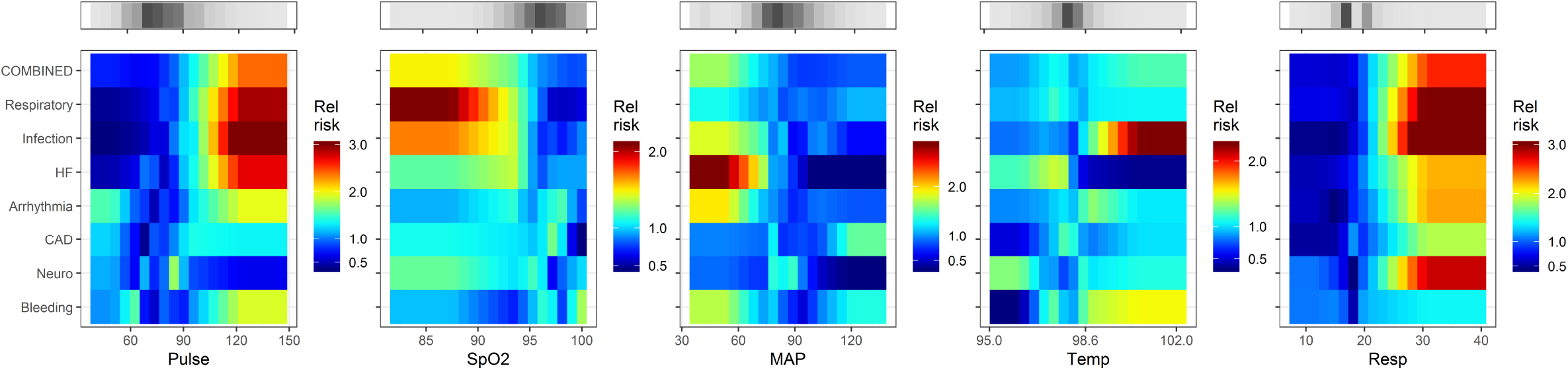
Heat maps that relate the relative risk of the reason for ICU transfer to the nurse-entered values of predictor vital signs. HR heart rate, RR respiratory rate, SpO2 oxygen saturation, MAP non-invasive mean arterial pressure.

The major finding of this subset of the overall results is that serum Na and the MAP, when low, signify impending ICU transfer for escalation of HF therapy, but they do not portend ICU transfer for any of the other reasons. Panel C shows that a rise in the continuously measured respiratory rate identifies patients with respiratory instability or infection to such a degree that ICU transfer takes place. These maps indicate the heterogeneity of patient characteristics, but are not quantitative tests. For that purpose, we used logistic regression models.

The top rows of the colored matrices show the risk profiles of a predictive model trained on all of the events combined. The smaller ranges of color demonstrate that the predictive utility of Na for HF, MAP for HF, and breaths from EKG for respiratory instability or infection are blunted. Thus, by combining outcomes, the ability of a multivariable statistical model to detect any individual clinical cause for ICU transfer is diminished.

Figures 3 to 5 show the complete catalog of relationship of risks to clinical causes for ICU transfer and measured variable. The finding is that the phenotypes of reasons for ICU transfer differ from each other. Consider bleeding (bottom row) as an example – it is uniquely marked by hypokalemia, hypocalcemia, thrombocytopenia and, of course, anemia. As a result, we would expect a predictive model trained to detect bleeding would not perform well in detecting other reasons for ICU transfer, nor would we expect models trained on other reasons for ICU transfer to do well at detecting bleeding.

### Statistical models

We limited the number of predictor variables to one per ten events. Vital signs figured in all 8 of the trained models; lab tests and continuous cardiorespiratory monitoring measures each figured in 6 models. The continuous cardiorespiratory monitoring measure most commonly included was EKG-derived respiratory rate (4 models). The CAD model had the most continuous cardiorespiratory monitoring measures, 2 of its 3 (detrended fluctuation analysis and coefficient of sample entropy) components.

The abilities of models to detect on- and off-target causes for ICU transfer are demonstrated in Figure 6, a matrix of ROC areas that result when the model targeting the event in the rows is used for early diagnosis of the event listed in the columns. The diagonal from bottom left to top right shows the ROC areas of the models as trained for the target events; off-diagonal values give the values for detecting off-target events. For example, the model to detect bleeding has a ROC area of 0.78 (the value in the cell at the lower left of the matrix), but that model has no ROC area of >0.6 in detecting other reasons for ICU transfer (bottom row), and the ROC areas of models trained for other events (left-hand column) exceed 0.6 in only case, the model trained on all events combined.

**Figure 5.**
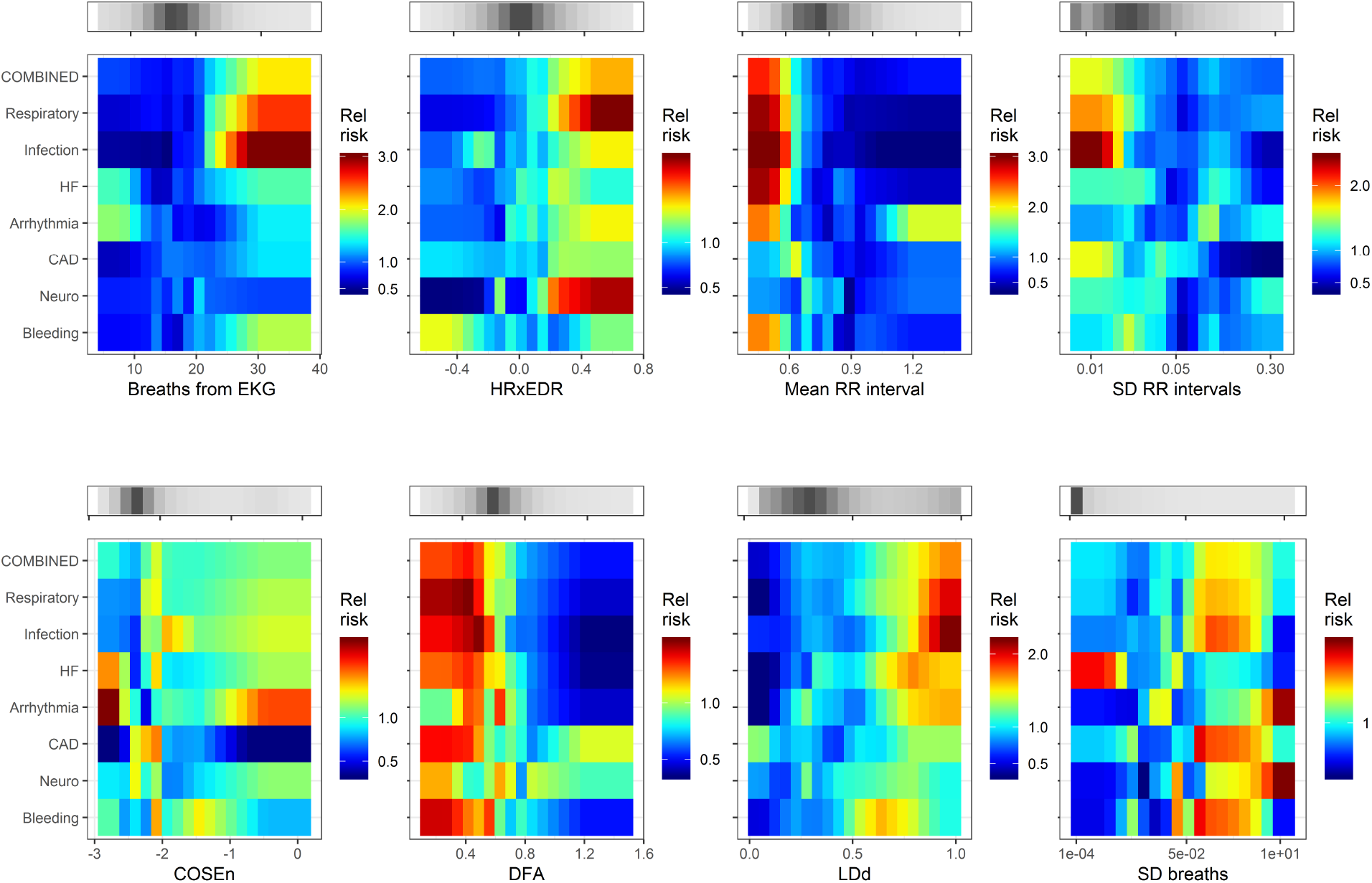
Heat maps that relate the relative risk of the reason for ICU transfer to measured values of predictors derived from cardiorespiratory monitoring. HRxEDR cross-correlation of heart rate and breathing rate derived from EKG; S.D. standard deviation, COSEn coefficient of sample entropy 56,57, DFA detrended fluctuation analysis 58, LDd local density score 39.

**Figure 6.**
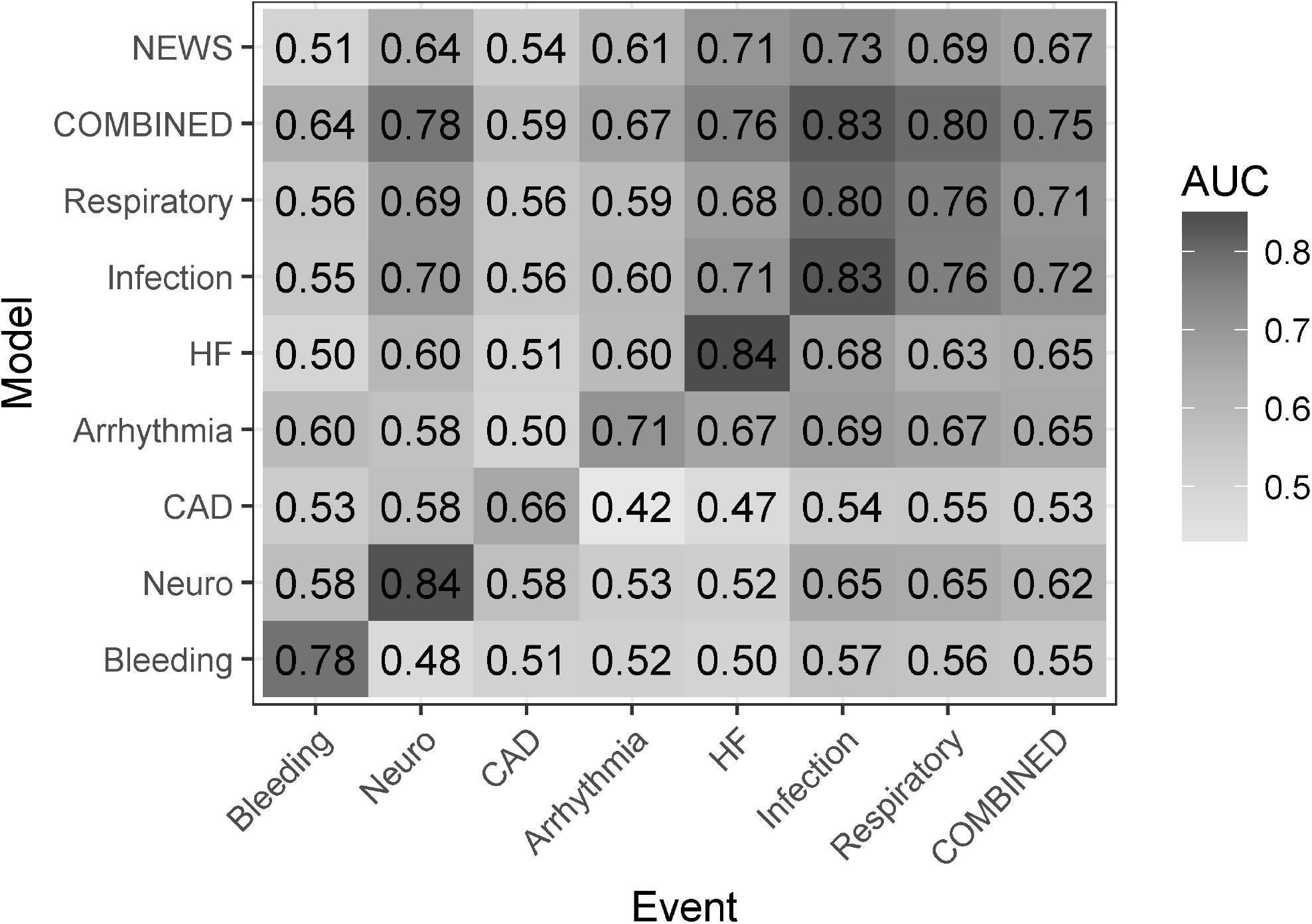
Receiver operating characteristic curve areas for models trained for the events listed in rows, and tested for events arranged as columns. The top row is the performance of the National Early Warning Score (NEWS); the next-to-top row and the final column represent results of a model trained on all the ICU transfer events combined. The gray scale reflects the value of the ROC area.

Additionally, Figure 6 shows variation of ROC areas from 0.66 to 0.84 along the diagonal, meaning some events are easier to detect than others. For examples, models to detect ICU transfer due to infection or neurological events have ROC area >0.8, while that for myocardial ischemia was <0.7. While reasonably common on this cardiac and cardiac surgery ward, we note that unstable coronary syndromes are not particularly good candidates for early detection. Arguably, there is no consistent prodrome for the instability of a coronary plaque.

We can directly compare the general model trained on all events to the models targeting specific reasons for ICU transfer by comparing the values in the second row (ROC area of the model targeting all events combined) to the values in the diagonal from lower left to upper right (ROC areas of the individual models). The combined event model performs about as well as the individually trained model infection (0.83 vs 0.83), and respiratory instability (0.80 vs 0.76). These are the most common reasons for ICU transfer (Figure 1), accounting for the finding.

The results of using the NEWS score to detect the individual or combined events is shown in the top row. The highest ROC area for NEWS was for infection, 0.73. Use of this single score for the entire population of combined events yielded ROC area 0.67. Of models trained for individual classes of ICU transfer, whose ROC areas appear in the diagonal, only that for unstable coronary symptoms was lower (0.66).

The mean NEWS ROC area was 0.63, significantly lower than the mean ROC area of 0.72 of the general model trained on combined events (*p*<0.01, paired t-test), which was lower than the mean ROC area of 0.78 of the specifically targeted models (*p*<0.05).

## Discussion

We studied statistical models for early detection of subacute potentially catastrophic illnesses leading to ICU transfer from a cardiac and cardiac surgery ward. This is a high-profile clinical event – the mortality rose by more than 40-fold in those who deteriorated ^39^. We targeted specific reasons for clinical deterioration, used predictors that were continuous rather than thresholded, and included mathematical analyses of continuous cardiorespiratory monitoring data. Our major findings are that phenotypes vary among the myriad of clinical conditions that lead to ICU transfer, and that statistical models trained specifically performed better than a model trained on all events combined, which in turn performed better than the untrained NEWS score.

We have used multivariable logistic regression models to test hypotheses about the difference in characteristics of endotypes of patients that deteriorated and required escalation to ICU care. The major hypothesis is that models trained on one endotype of patient do not necessarily perform well in detecting other kinds of patients. This directly tests the prevailing practice of using a single predictive model throughout the hospital. It is important to note that we here use the statistical models only for hypothesis testing and not for their clinical impact. We have not tested these models in a clinical setting, and a reasonable question is how to implement so many. We favor the idea of calculating all the model results for all the patients all the time, and presenting the clinician with a summary statistic. We tested ways to combine model results in the setting of early detection of sepsis in the neonatal ICU, where we found that the most pessimistic model was the most accurate. Other ways such as the ensemble mean or median might be equally or more useful.

It is a natural exercise of clinicians to synthesize disparate data elements into a clinical picture of the patient. After 1981 when Knaus, Wagner and colleagues introduced the APACHE (Acute Physiology and Chronic Health Evaluation) score, more methods have been introduced that combine elements of clinical data to yield quantitative estimates of patients’ health status and risk.^48^ While varied in inputs, targets, patient populations, and mathematical tools, most are single scores meant for universal use. ^34^ This is not altogether unreasonable. For example, we found that large abrupt spikes in risk estimation using a single model to identify patients at risk for ICU transfer had a positive predictive value 25% for imminent acute adverse event.^40^ But use of only a single model is a limited approach, as there are many paths of deterioration – accurate capture of all of these paths at the same time is challenging.

The need to look for multiple modes of critical illness was recently supported by the finding of four sepsis phenotypes of responses and outcomes.^49^ We found different sepsis signatures in the medical as opposed to the surgical ICU.^38^ Thus, no single predictive model for sepsis seems adequate. Likewise, there are distinct respiratory deterioration phenotypes within COPD and acute respiratory distress syndrome.^50,51^ This heterogeneity of manifestations of illness, a point that resonates with bedside clinicians, argues against models that use the same thresholds for all situations. (An exception is sepsis in premature infants, where heart rate characteristics index monitoring identifies an abnormal phenotype that is common to many acute neonatal illnesses,^52^ and in a large RCT, was found to save lives.^53^)

We emphasize the importance of continuous cardiorespiratory monitoring in early detection of subacute potentially catastrophic illnesses. We found distinct physiological signatures for sepsis, hemorrhage leading to large unplanned transfusion and respiratory failure leading to urgent unplanned intubation in the adult Medical and Surgical ICUs. Display of risk estimates using two of the resulting statistical models based solely on cardiorespiratory monitoring ^38^ was associated with a 50% reduction in the rate of septic shock.^54^ Moreover, cardiorespiratory monitoring data add to vital signs and lab tests in the population reported here.^39^ We propose that no scheme that omits cardiorespiratory monitoring data will perform as well as those that include it.

A strength of this study is the individual review of charts to identify reasons for ICU transfer. Clinicians recognize that illness presentations are complex, nuanced, and not well-documented.^55^ Statistical models trained for one kind of deterioration had poor performance in detecting other kinds. As examples, bleeding – a common form of deterioration on our mixed medical-surgical cardiac ward – was well-detected by a model trained specifically for it, but no other model had reasonable performance; hemoglobin was, logically, a powerful prediction of ICU transfer for bleeding but not at all useful when myocardial ischemia was the problem; coefficient of sample entropy, a detector of atrial fibrillation, was useful only for identifying patients transferred to the ICU for arrhythmia but nothing else.

There are other approaches. For example, Redfern and colleagues reported an early warning system for ICU transfer based on vital signs alone.^10^ Their approach differed from ours in several key ways. First, there was no clinical review of the reasons that patients deteriorated and required escalation to ICU care. We found that fully 50% of ICU transfers were not due to clinical deterioration^38^. Addition of a large number of non-relevant cases negatively impacts the relevance of predictive models that result by inflating confidence intervals. Second, they studied only vital signs and lab tests. They also reported the large proportion of late or missed vital sign measurements – a median of 44%, presumably from the same time period and from one of the study hospitals. Third, bright cut-offs were used, thus a patient who, on successive measurements, had an increase in respiratory rate from 12 to 20 breaths per minute, a decrease in O_2_ saturation from 100% to 96%, an increase in temperature from 36.2 to 38.0, a fall in BP from 218 to 112 mmHg, an increase in heart rate from 51 to 90 beats per minute – all clinically worrisome when simultaneous – has an unchanging NEWS score of 0. Finally, they made no distinction among the clinical reasons for transfer.

Here, we show large differences in signatures of illness and performance of statistical models in early identification of patients at risk of ICU transfer. The approach of Redfern and colleagues is exemplary of most efforts in clinical surveillance for early detection of subacute potentially catastrophic illnesses. We propose more precise methods: clinician review of events so that predictive models are more focused, addition of continuous cardiorespiratory monitoring when it is available to fill in the timeline between nurse visits and blood draws, and the use of predictive models that individually target the diagnoses that lead to ICU transfer in our patients.

Our results argue against a one-size-fits-all approach, no matter how Big the Data nor how Deep the Learning. In this era of increasingly precise and personalized medicine, we propose *precision predictive analytics monitoring* as a more focused and clinically informed approach to early detection of subacute potentially catastrophic illnesses in the hospitalized patient.

## Data Availability

Clinical data obtained with IRB approval for the 8111 patients in this study is stored under encryption on servers at the University of Virginia. Inquiries regarding access to data from the University of Virginia's Center for Advanced Medical Analytics should be done by contacting J Randall Moorman, M.D. via email at.

